# Personal care product use and risk of adult-onset asthma: findings from the Sister Study

**DOI:** 10.1101/2025.01.10.25320341

**Authors:** Jungeun Lim, Che-Jung Chang, Alexandra J. White, Shelton Lo, Hantao Wang, Gabriel Goodney, Rui Miao, Amisha V. Barochia, Véronique L. Roger, Dale P. Sandler, Jason Y.Y. Wong

## Abstract

**BACKGROUND:** Personal care products (PCPs) contain endocrine-disrupting chemicals (EDCs) linked to hormonally-sensitive diseases. Population studies have found associations between prenatal EDC exposure and childhood asthma; however, few have investigated adult-onset asthma.

**OBJECTIVES:** We investigated the associations between commonly used PCPs and the risk of adult-onset asthma in a prospective cohort study of U.S. women.

**METHODS:** We analyzed 39,408 participants from the Sister Study (2003-2009). The participants self-reported their usage frequency of 41 PCPs in the 12-month period before baseline. Latent classes were used to identify groups with similar usage patterns (‘infrequent’, ‘moderate’, ‘frequent’) within types of products (‘beauty’, ‘everyday hair’, ‘hygiene’, and ‘skincare’). Multivariable Cox regression models were used to assess the associations between PCP use and incident adult-onset asthma.

**RESULTS:** Over an average 12.5-year follow-up, 1,774 incident asthma cases were identified. Compared to infrequent users, moderate (hazard ratio [HR]=1.21 (95% confidence interval (CI):1.07,1.37)) and frequent (HR=1.22 (95%CI:1.08,1.38)) users of beauty products had significantly higher asthma risk. Similar associations were observed for hygiene (moderate: HR=1.14 (95%CI:1.01,1.29) and frequent: HR=1.20 (95%CI:1.06,1.36)) and skincare products (moderate: HR=1.21 (95%CI:1.06,1.38) and frequent: HR=1.20 (95%CI:1.06,1.35)). Several individual everyday hair products (hair spray, hair styling gel/mousse, and pomade or hair grease) were positively associated with asthma risk, but associations were not detected for everyday hair latent classes.

**DISCUSSION:** Our findings suggest that PCP use potentially contributes to future risk of adult-onset asthma among women. These novel findings reinforce the need for regulation of PCPs and their components to reduce the burden of asthma.

## Introduction

Personal care products (PCPs) contain complex mixtures of endocrine-disrupting chemicals (EDCs) including per- and polyfluoroalkyl substances (PFAS), parabens, phthalates, phenols ^1–3^, as well as other hazardous components. Chronic exposure to EDCs has been facilitated by the widespread use of PCPs in the daily lives of the general population ^4^. The patterns of PCP use differ by subgroups defined by sex ^5^, race/ethnicity ^3,6^, and age ^7^. In particular, women have been found to use disproportionately more PCPs compared to men and may be at higher risk of adverse health consequences of exposure. EDCs found in PCPs have been linked to hormonally-sensitive chronic diseases, including cardiovascular diseases as well as breast and ovarian cancer among women ^8,9^.

In the United States (U.S.), asthma is one of the most common chronic, non-communicable respiratory diseases, affecting over 27 million people ^10,11^ and the prevalence among adults increased from 6.9% in 2001 to 8.0% in 2021 ^12^. Asthma is more common among adult women (10.8%) than men (6.5%); however, the underlying reasons are unclear ^13^. Recently, there has been growing concern about the impact of environmental chemicals on asthma development. Asthma is a hormonally-response disease ^14–16^ and population studies have reported potential associations with exposure to EDCs in PCPs ^17,18^. However, most of the current literature focuses on the risk of childhood asthma, while few epidemiological studies have investigated the association between exposure to EDCs and the risk of adult-onset asthma.

To address the gaps in knowledge, we investigated the associations between PCP use and risk of incident adult-onset asthma among U.S. women from the Sister Study. To better capture the risks associated with the simultaneous use of multiple products in real-world settings, we investigated the effects of individual personal care products (PCPs) and groups of PCPs. Further, given previous evidence suggesting heterogeneity by subgroups, we explored whether these relationships vary by menopausal status and race/ethnicity.

## Methods

### Study design and population

The Sister Study is a prospective cohort study that enrolled 50,884 women residing in U.S. and Puerto Rico from 2003 to 2009 ^19^. Women were eligible for the Sister Study if they were 35-74 years of age, had at least one sister diagnosed with breast cancer, and never had a diagnosis of ductal carcinoma *in situ* (DCIS) or invasive breast cancer. Women completed a computer-assisted telephone interview and self-administered questionnaires at enrollment to assess demographics (e.g., age, race/ethnicity, educational attainment, and household income), reproductive and medical history (e.g., menopausal status and family history of asthma), and lifestyle factors (e.g., smoking status, alcohol consumption, and recreational physical activity). Height and weight were measured by a trained examiner during a home visit at baseline and used to determine body mass index (BMI, kg/m^2^). Detailed information on data collection and questionnaires can be found on the Sister Study website ^20^.

Among the 50,884 women, we excluded those who were diagnosed with prevalent asthma prior to baseline (n=7,179), diagnosed at unknown timing relative to the completion of all baseline study components (n=1,613), had uncertain asthma diagnoses (n=5), or withdrew from the study (n=5). We also excluded women who did not complete the PCP questionnaire module (n=2,674). The final analytic sample included 39,408 women. To reduce the impact of undiagnosed patients with prevalent asthma, we additionally excluded subjects who reported asthma symptoms (both wheeze and chronic cough at baseline, n=134) in sensitivity analyses.

The Sister Study was approved by the institutional review boards at the National Institute Health. Data for the current analysis included person-time through September 30, 2021 (Data Release 11.1). All study participants provided written informed consent.

### Personal care product use

The participants self-reported their frequency of use of 41 PCPs in the 12-month period before baseline. The frequency of use for each PCP included: 1) did not use, 2) used less than once a month, 3) used 1-3 times per month, 4) used 1-5 times per week, 5) used more than 5 times per week. To identify latent classes of PCP use, the 41 PCPs were grouped into twelve beauty products (PCPs listed in Table S3), seven everyday hair products (PCPs listed in Table S4), eight hygiene products (PCPs listed in Table S5), and fourteen skincare products (PCPs listed in Table S6) based on a priori knowledge and correlation structure ^9^.

### Incident adult-onset asthma

Participants were contacted for either annual health update forms or detailed follow-up questionnaires, which included health updates on new doctor’s diagnosis of asthma. Participants who reported a diagnosis of asthma during the follow-up were defined as incident cases (n=1,774). Among asthma cases, 98.5% self-reported a diagnosis of unspecified asthma (n=1,747), and the remaining 1.6% self-reported a diagnosis of mild asthma (n=6), severe asthma (n=8), eosinophilic asthma (n=4), cough-variant asthma (n=4), exercise-induced asthma (n=2), or unspecified asthma with acute exacerbation (n=3).

### Statistical analysis

The distribution of the participants’ baseline characteristics was presented by adult-onset asthma. We used Spearman’s rank correlation coefficients to estimate pairwise correlations for frequency of use for 41 PCPs.

Multivariable Cox proportional hazards regression models were used to estimate hazard ratios (HRs) and 95% confidence intervals (CIs) for the associations between individual PCPs and the adult-onset asthma risk. Consistent with previous studies, we accounted for age as the timescale in the Cox models ^9,21^. Further, we adjusted for race/ethnicity (African American/Black, Hispanic/Latina non-Black, non-Hispanic White, other, or missing), educational attainment (high school or less, some college, college and above, or missing), annual household income (<$50,000, $50,000-<$100,000, ≥$100,000, or missing), menopausal status at enrollment (premenopausal, postmenopausal, or missing), smoking status (never, past, current, or missing), secondhand smoke exposure (continuous, years), alcohol consumption (never or past, current <1 drink, current ≥1 drinks, or missing), and BMI (continuous, kg/m^2^).

We used PROC LCA in SAS statistical software (version 9.4; SAS Institute Inc.) to conduct latent class analysis (LCA) to identify groups of individuals with similar patterns of PCP use ^22^. Five LCA models were estimated for each product group (i.e., beauty products, everyday hair products, hygiene products, and skincare products) with two to six latent classes. We used the Akaike’s information criterion (AIC), the Bayesian information criterion (BIC), G^2^ statistics, and entropy to identify the best fit model and select the optimum number of classes as previously described in detail ^23^. The latent classes were labeled on an ordered scale according to frequency of use (e.g., ‘infrequent’, ‘moderate’, and ‘frequent’). The associations between latent classes and the adult-onset asthma risk were assessed using multivariable Cox proportional hazards regression models with the same covariate adjustment mentioned above.

We conducted additional stratified analyses by race/ethnicity and menopausal status at baseline. Interactions were assessed through likelihood ratio tests by comparisons of Cox models with and without the cross-product of each factor (race/ethnicity and menopausal status) and latent class. P-values for trend across ordered latent classes were calculated.

## Results

### Study population characteristics

Among 39,408 participants, 8.6% self-reported their race/ethnicity as Black, 84.4% as non-Hispanic White, and less than 7% as Hispanic or other (Table 1). Approximately two-thirds of the participants (66.7%) were post-menopausal women. During a mean follow-up of 12.5 years, 1,774 women reported diagnosis with incident adult-onset asthma. Compared with people without asthma diagnosis, those with asthma were more likely to have had a higher BMI, drink less alcohol, have a lower annual household income, have greater exposure to secondhand smoke, and use electronic cigarettes. They were also more likely to be post-menopausal or have a family history of asthma.

**Table 1.**
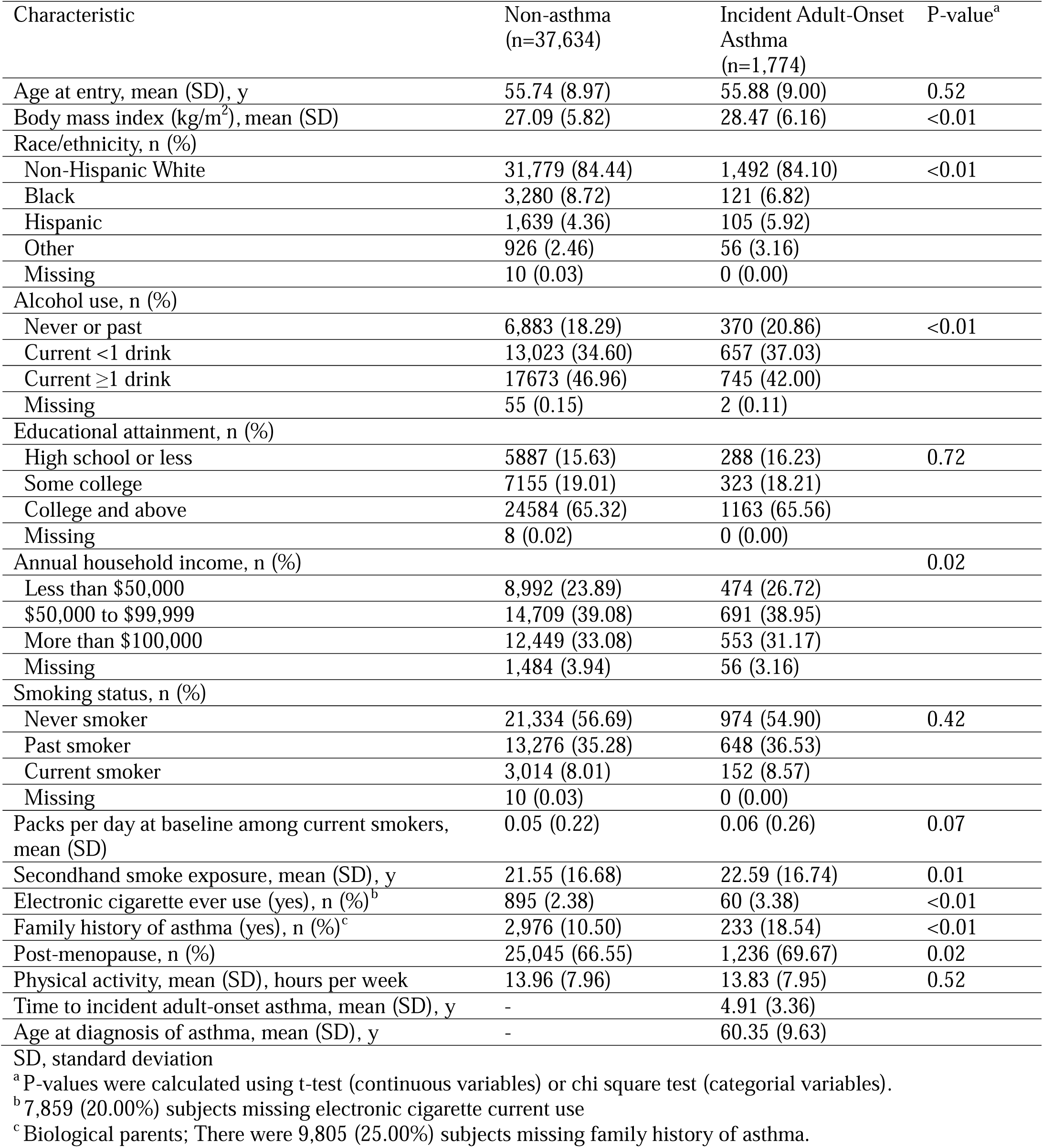
Characteristics of Sister Study participants according to asthma status (N=39,408)

| Characteristic | Non-asthma<br>(n=37,634) | Incident Adult-Onset<br>Asthma<br>(n=1,774) | P-value<sup>a</sup> |
| --- | --- | --- | --- |
| Age at entry, mean (SD), y | 55.74 (8.97) | 55.88 (9.00) | 0.52 |
| Body mass index (kg/m<sup>2</sup>), mean (SD) | 27.09 (5.82) | 28.47 (6.16) | <0.01 |
| Race/ethnicity, n (%) |  |  |  |
| Non-Hispanic White | 31,779 (84.44) | 1,492 (84.10) | <0.01 |
| Black | 3,280 (8.72) | 121 (6.82) |  |
| Hispanic | 1,639 (4.36) | 105 (5.92) |  |
| Other | 926 (2.46) | 56 (3.16) |  |
| Missing | 10 (0.03) | 0 (0.00) |  |
| Alcohol use, n (%) |  |  |  |
| Never or past | 6,883 (18.29) | 370 (20.86) | <0.01 |
| Current <1 drink | 13,023 (34.60) | 657 (37.03) |  |
| Current ≥1 drink | 17,673 (46.96) | 745 (42.00) |  |
| Missing | 55 (0.15) | 2 (0.11) |  |
| Educational attainment, n (%) |  |  |  |
| High school or less | 5,887 (15.63) | 288 (16.23) | 0.72 |
| Some college | 7,155 (19.01) | 323 (18.21) |  |
| College and above | 24,584 (65.32) | 1,163 (65.56) |  |
| Missing | 8 (0.02) | 0 (0.00) |  |
| Annual household income, n (%) |  |  | 0.02 |
| Less than \$50,000 | 8,992 (23.89) | 474 (26.72) | |
| \$50,000 to \$99,999 | 14,709 (39.08) | 691 (38.95) | |
| More than \$100,000 | 12,449 (33.08) | 553 (31.17) | |
| Missing | 1,484 (3.94) | 56 (3.16) |  |
| Smoking status, n (%) |  |  |  |
| Never smoker | 21,334 (56.69) | 974 (54.90) | 0.42 |
| Past smoker | 13,276 (35.28) | 648 (36.53) |  |
| Current smoker | 3,014 (8.01) | 152 (8.57) |  |
| Missing | 10 (0.03) | 0 (0.00) |  |
| Packs per day at baseline among current smokers, mean (SD) | 0.05 (0.22) | 0.06 (0.26) | 0.07 |
| Secondhand smoke exposure, mean (SD), y | 21.55 (16.68) | 22.59 (16.74) | 0.01 |
| Electronic cigarette ever use (yes), n (%)<sup>b</sup> | 895 (2.38) | 60 (3.38) | <0.01 |
| Family history of asthma (yes), n (%)<sup>c</sup> | 2,976 (10.50) | 233 (18.54) | <0.01 |
| Post-menopause, n (%) | 25,045 (66.55) | 1,236 (69.67) | 0.02 |
| Physical activity, mean (SD), hours per week | 13.96 (7.96) | 13.83 (7.95) | 0.52 |
| Time to incident adult-onset asthma, mean (SD), y | - | 4.91 (3.36) |  |
| Age at diagnosis of asthma, mean (SD), y | - | 60.35 (9.63) |  |

SD, standard deviation
<sup>a</sup> P-values were calculated using t-test (continuous variables) or chi square test (categorical variables).
<sup>b</sup> 7,859 (20.00%) subjects missing electronic cigarette current use
<sup>c</sup> Biological parents; There were 9,805 (25.00%) subjects missing family history of asthma.

The Spearman correlation coefficient matrix between frequency of PCP use is presented in Figure 1. Spearman correlation coefficients (r_s_) ranged from 0.20 to 0.56 for mascara, eyeshadow, eyeliner, cleansing cream, foundation, blush, makeup remover, face cream, and anti-aging product. The highest correlation was observed between frequency of nail polish use and frequency of nail polish remover use (r_s_=0.91). The frequency of use of the three types of talc showed a moderate correlation with each other (r_s_=0.34–0.50). Several hair products (pomade and hair food) were inversely correlated with mascara, blush, face cream, anti-aging product, self-tanning product, shampoo, conditioner, hair spray, and hair styling gel.

**Figure 1:**
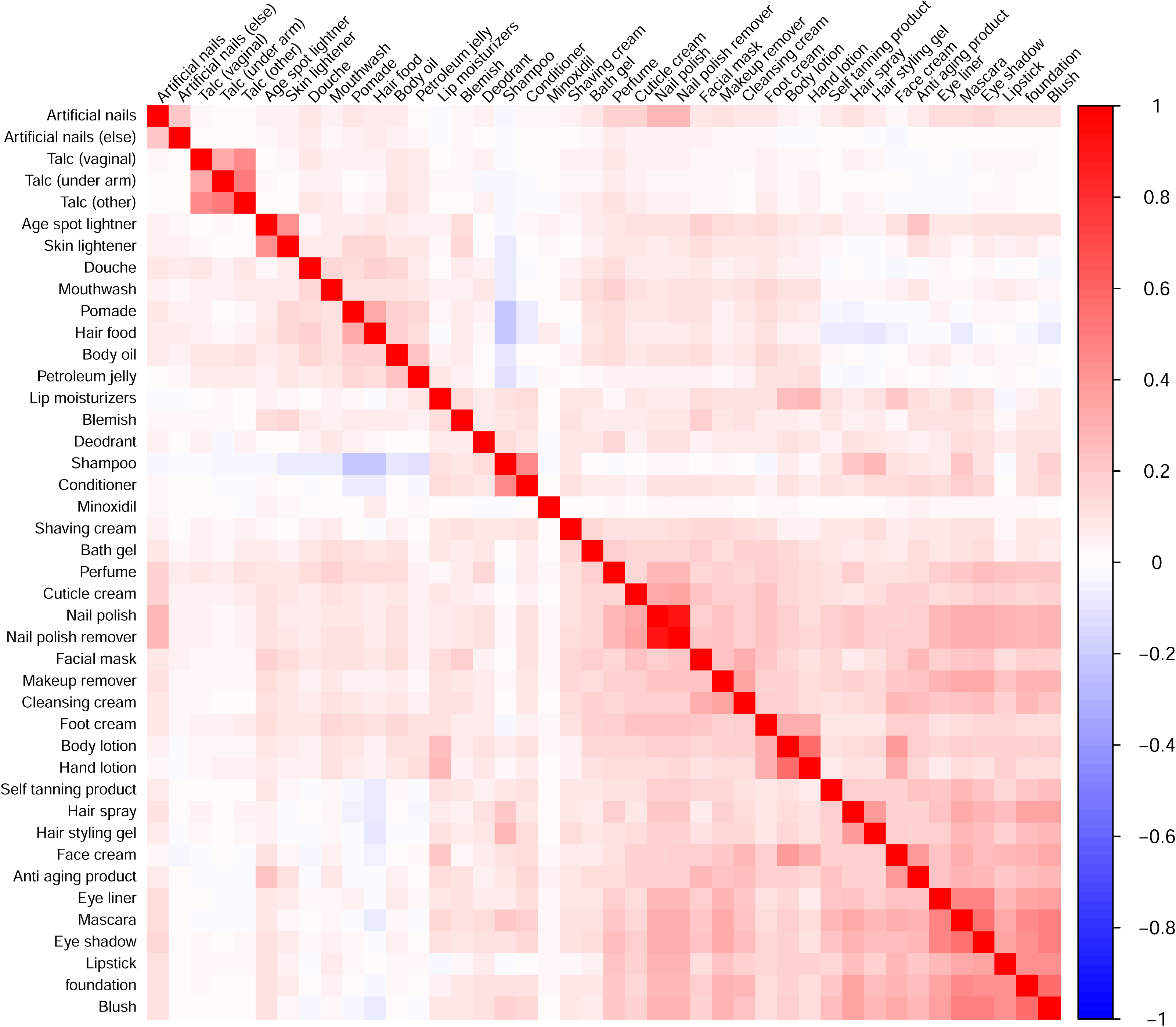
Spearman correlation coefficient matrix for frequency of personal care product use.

### Latent Classes of PCP Use and Incident Adult-onset Asthma

In the latent class analysis, our final model included three latent classes for each product category as described in detail in Table S1 in the online supplement. We found that moderate and frequent users of beauty products had significantly higher risk of adult-onset asthma compared to infrequent users, even after adjusting for important confounders (moderate users, HR=1.21 (95%CI:1.07,1.37) and frequent users, HR=1.22 (95%CI:1.08,1.38); P-trend<0.01) (Figure 2 and Table 2). Similar positive associations were observed for hygiene (moderate users, HR=1.14 (95%CI:1.01,1.29) and frequent users, HR=1.20 (95%CI:1.06,1.36); P-trend<0.01) and skincare products (moderate users, HR=1.21 (95%CI:1.06,1.38) and frequent users, HR=1.20 (95%CI:1.06,1.35); P-trend =0.01). Conversely, latent classes of everyday hair product use were not associated with asthma risk (Table 2). In a sensitivity analysis excluding people with asthma symptoms at baseline, the results were consistent with the main analyses (Table S2).

**Figure 2.**
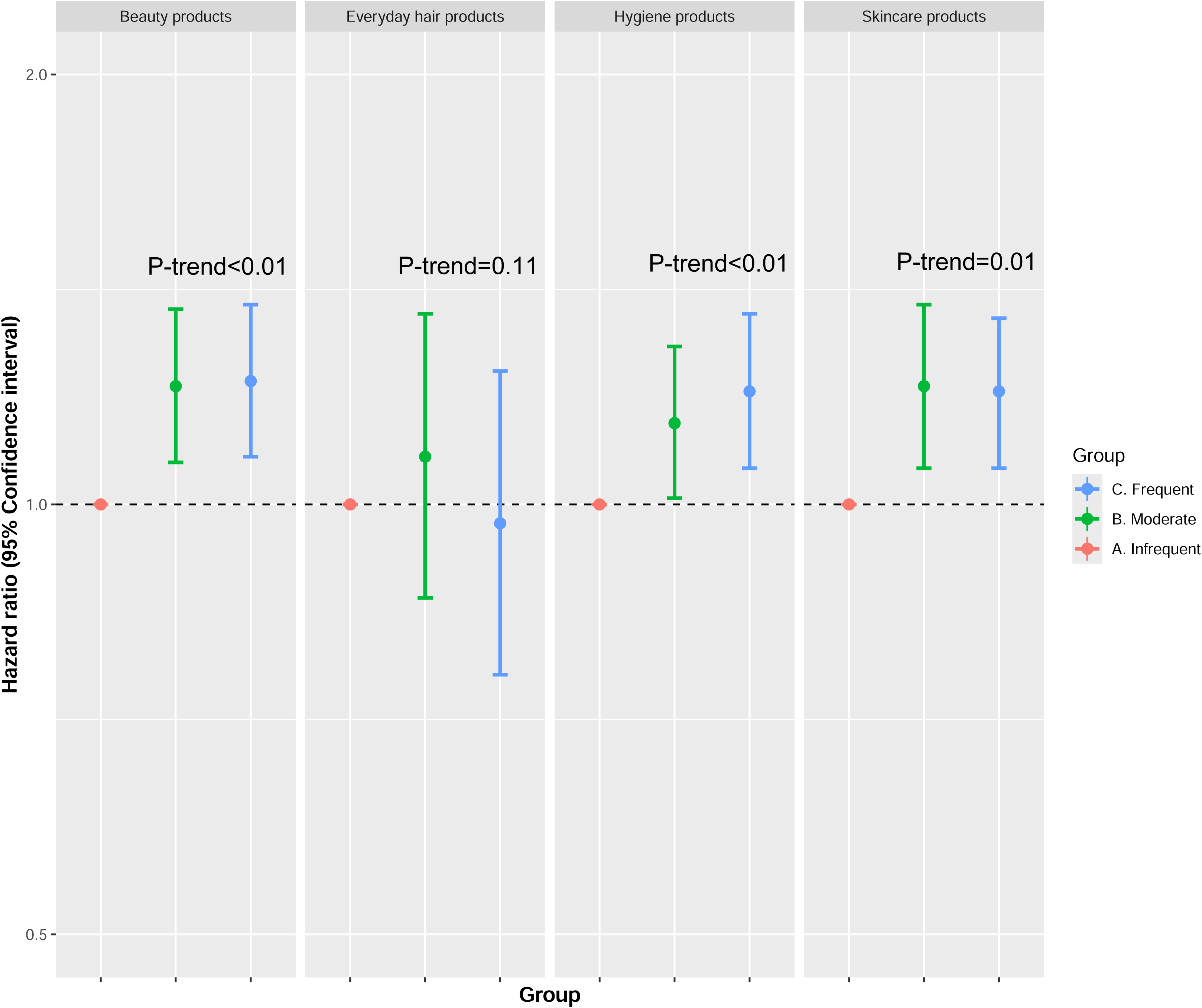
Hazard ratios (HR) and 95% confidence intervals (CI) for the association between personal care product latent classes and asthma risk. Models accounted for age by using age as the timescale, where participants entered the analysis at their enrollment age (left-truncation) and accrued person-time until they exited at their asthma diagnosis or were administratively censored at their age at last follow-up. In addition, models were adjusted for race and ethnicity (African American/Black, Hispanic/Latina non-Black, non-Hispanic White, other, or missing), education level (high school or less, some college, college and above, or missing), annual household income (<$50,000, $50,000-<$100,000, ≥$100,000), menopausal status at enrollment (premenopausal, postmenopausal, or missing), smoking status (never, past, current, or missing), secondhand smoke exposure (continuous, y), alcohol consumption (never or past, current <1 drink, current ≥1 drinks, or missing), and body mass index (continuous, kg/m^2^). The number of cases and the total number of subjects (Case/n) for each class of beauty products were: 458/11,235, 607/12,791, and 709/15,382. Case/n for each class of everyday hair products were: 138/3,660, 981/21,075, and 653/14,647. Case/n for each class of hygiene products were: 547/14,664, 718/14,929, and 508/9,798. Case/n for each class of skincare products are: 459/11,030, 504/10,154, and 811/18,223.

**Table 2.**
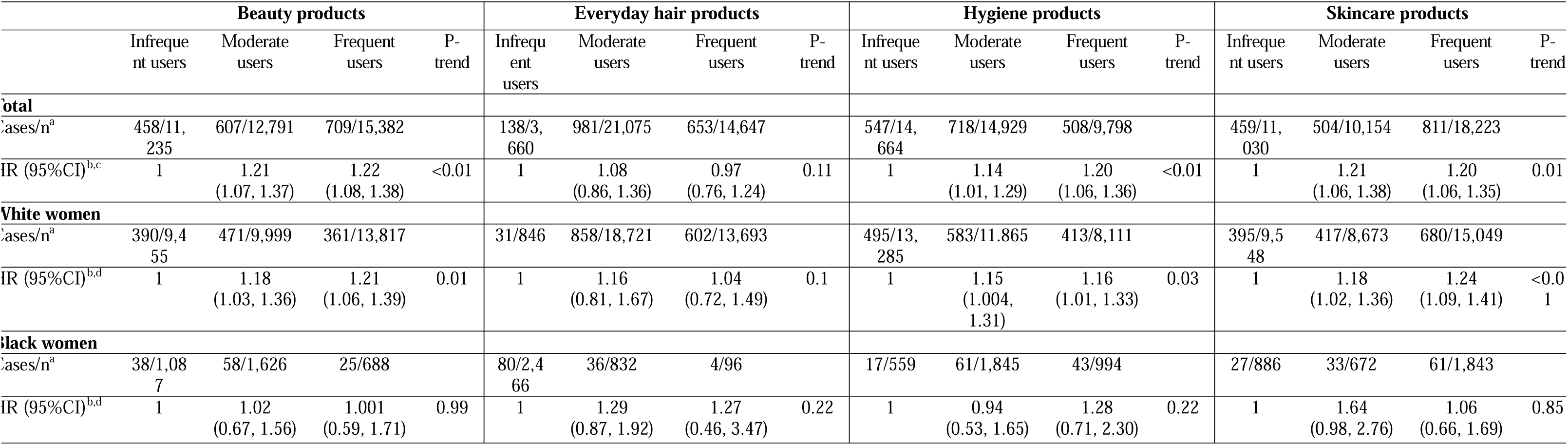
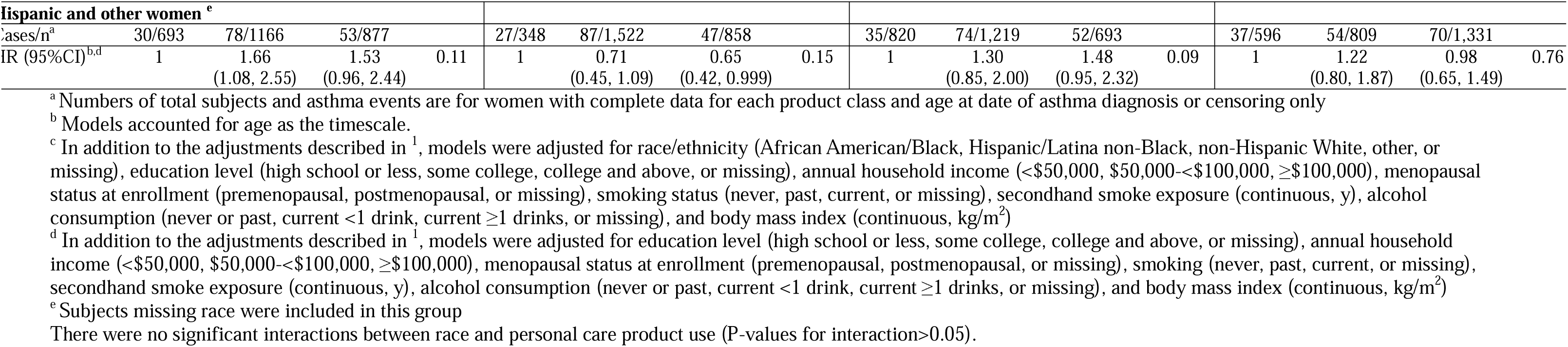
Hazard ratios (HR) and 95% confidence intervals (CI) for the association between personal care product latent classes and adult-onset asthma risk by race.

We found some evidence of heterogeneity in the main findings by race/ethnicity; however, there were no significant interactions between race/ethnicity and PCP use on asthma risk (P for interaction>0.05). Among non-Hispanic White women, latent groups that used beauty (moderate users, HR=1.18 (95%CI:1.03,1.36) and frequent users, HR=1.21 (95%CI:1.06,1.39); P-trend=0.01), hygiene (moderate users, HR=1.15 (95%CI:1.004,1.31) and frequent users, HR=1.16 (95%CI:1.01,1.33); P-trend=0.03), or skincare products (moderate users, HR=1.18 (95%CI:1.02,1.36) and frequent users, HR=1.24 (95%CI:1.09,1.41); P-trend<0.01) more frequently had a higher risk of asthma compared to infrequent users. Among Black women, moderate use of skincare (HR=1.64 (95%CI:0.98,2.76)) or everyday hair (HR=1.29 (95%CI:0.87,1.92)) and the frequent use of hygiene products (HR=1.28 (95%CI:0.71,2.30)) were associated with non-significant increased risk of asthma. In Hispanic and other racial women, moderate users of beauty products had increased risk of asthma relative to infrequent users (HR=1.66 (95%CI:1.08, 2.55); Table 2). Frequent use of hygiene products was associated with increased risk of asthma in Hispanic and other racial women, at borderline significance (HR=1.48 (95%CI:0.95, 2.32)).

In analyses stratified by menopausal status, premenopausal women who used beauty products frequently had significantly higher asthma risk compared to infrequent users (moderate users, HR=1.35 (95%CI:1.06,1.73) and frequent users, HR=1.47 (95%CI:1.15,1.87); P-trend<0.01) (Table 3). In postmenopausal women, latent groups that used hygiene (P-trend=0.02), or skincare products (P-trend=0.03) more frequently had a higher risk of asthma compared to infrequent users. However, there were no significant interactions between menopausal status and PCP use on asthma risk (P for interaction>0.05).

**Table 3.** Hazard ratios (HR) and 95% confidence intervals (CI) for the association between personal care product latent classes and adult-onset asthma risk by menopausal status.

|  | **Premenopausal women** |  | **Postmenopausal women** |  |
| --- | --- | --- | --- | --- |
| Product category/class | Cases/n<sup>a</sup> | HR (95%CI)<sup>b</sup> | Cases/n<sup>a</sup> | HR (95%CI)<sup>b</sup> |
| **Beauty products** |  |  |  |  |
| A. Infrequent users | 99/3,146 | 1 | 359/8,089 | 1 |
| B. Moderate users | 198/4,607 | 1.35 (1.06, 1.73) | 409/8,181 | 1.17 (1.01, 1.35) |
| C. Frequent users | 241/5,370 | 1.47 (1.15, 1.87) | 468/10,011 | 1.13 (0.98, 1.31) |
| P-trend |  | <0.01 |  | 0.11 |
| **Everyday hair products** |  |  |  |  |
| A. Infrequent users | 33/1,109 | 1 | 105/2,549 | 1 |
| B. Moderate users | 203/5,185 | 1.21 (0.73, 2.03) | 778/15,889 | 1.07 (0.83, 1.38) |
| C. Frequent users | 301/6,817 | 1.28 (0.76, 2.17) | 352/7,829 | 0.88 (0.67, 1.16) |
| P-trend |  | 0.39 |  | 0.02 |
| **Hygiene products** |  |  |  |  |
| A. Infrequent users | 91/2,923 | 1 | 456/11,740 | 1 |
| B. Moderate users | 317/7,419 | 1.17 (0.92, 1.49) | 401/7,507 | 1.14 (0.99, 1.32) |
| C. Frequent users (talcum powder users) | 130/2,777 | 1.26 (0.96, 1.67) | 378/7,021 | 1.18 (1.02, 1.36) |
| P-trend |  | 0.11 |  | 0.02 |
| **Skincare products** |  |  |  |  |
| A. Infrequent users | 102/2,852 | 1 | 357/8,178 | 1 |
| B. Moderate users | 186/4,142 | 1.23 (0.96, 1.57) | 318/6,010 | 1.21 (1.03, 1.41) |
| C. Frequent users | 250/6,129 | 1.26 (0.99, 1.60) | 561/12,092 | 1.18 (1.02, 1.35) |
| P-trend |  | 0.08 |  | 0.03 |

<sup>a</sup> Numbers of total subjects and asthma events are for women with complete data for each product class and age at date of asthma diagnosis or censoring only
<sup>b</sup> Models accounted for age as the timescale. In addition, models were adjusted for race/ethnicity (African American/Black, Hispanic/Latina non-Black, non-Hispanic White, other, or missing), education level (high school or less, some college, college and above, or missing), annual household income (<\$50,000, \$50,000-<\$100,000, ≥\$100,000), smoking status (never, past, current, or missing), secondhand smoke exposure (continuous, y), alcohol consumption (never or past, current <1 drink, current ≥1 drinks, or missing), and body mass index (continuous, kg/m<sup>2</sup>)
There were no significant interactions between menopausal status and personal care product use (P-values for interaction>0.05).

### Use of Individual Products and Incident Adult-onset Asthma

Among the 12 individual beauty products, frequent users of blush or rouge (P-trend<0.01), lipstick (P-trend=0.01), artificial nails or fill-ins (P-trend=0.03), and cuticle cream (P-trend<0.01) had higher risk of asthma compared with non-users (Table S3). A higher risk of asthma was observed in the group that used eye liner, eye shadow, or foundation makeup compared to non-users, but the trend was non-monotonic. For the individual everyday hair products, frequent users of pomade or hair grease had increased risk of asthma compared with non-users (P-trend<0.01). The groups that used hair spray or hair styling gel/mousse 1-5 times per week had a higher risk of asthma than the non-users, but the trend was non-monotonic (Table S4). Among the hygiene products, only talcum powder use on the vaginal area had a significant positive association with asthma risk (P-trend=0.02, Table S5). Among the 14 individual skincare products, anti-aging/wrinkle products use and age spot lightener use was positively associated with asthma risk (P-trend<0.01). Compared to non-users, increased risks of asthma were found in the groups who used blemish/acne products, cleansing cream, facial masks, or self-tanning products, but the trends were not monotonic (Table S6).

## Discussion

In this large prospective cohort study with detailed information on use of PCPs, we found that frequent use of beauty, hygiene, and skincare products was associated with increased risk of adult-onset asthma among U.S. women. Several individual everyday hair products were positively associated with adult-onset asthma risk, but the latent classes for everyday hair products were not. To our knowledge, this is the first study to investigate the relationships of commonly used individual and complex mixtures of PCPs with future risk of adult-onset asthma.

Previous epidemiologic studies that investigated the impact of exposure to EDCs on the risk of asthma have primarily focused on the association between prenatal exposure to EDCs and their children’s risk of childhood asthma. Data from a Danish cohort of 738 pregnant women and their children showed that higher maternal perfluorooctane sulfonate (PFOS) and perfluorooctanoate (PFOA) exposure during pregnancy was associated with increased risk of non-atopic asthma phenotype by age 6 years ^24^. Berger et al. measured urinary concentration of the 10 PCP biomarkers (three phthalate metabolites, three parabens, and four other phenols), and suggested that prenatal urinary concentration of triclosan was associated with increased odds of probable asthma in their children, especially in girls ^25^. Prenatal urinary metabolites of short-chain phthalates such as butylbenzyl phthalate (BBzP) and di-n-butyl phthalate (DnBP) were found to be positively associated with a history of asthma-like symptoms and the diagnosis of current asthma in children ^26^. Data from the National Health and Nutrition Examination Survey (NHANES) 2005–2006 showed that higher urinary monobenzyl phthalate (MBzP) concentration was associated with current asthma and allergic symptoms in adults but not in children ^27^. However, there are few prospective cohort studies on adult-onset asthma risk, and most investigations assessed single EDC at a time. Using one spot serum or urine biomarker measurements may have limitations in reflecting exposure to mixtures of multiple chemicals, especially non-persistent chemicals ^28^.

Several studies have suggested that the differences in the use of PCPs may contribute to the racial/ethnic disparities in asthma risk ^29^. Hair products and feminine hygiene products used by Black women contained multiple chemicals associated with asthma, and higher levels of paraben and diethyl phthalate (DEP) were observed in biomonitoring samples from Black women compared with non-Hispanic White women ^29–31^. In our study, the frequency of beauty, hygiene, and skincare product use were positively associated with asthma risk in non-Hispanic White women. Stronger association between the frequent use of hygiene products and increased risk of asthma was observed in Black, Hispanic, and other minority groups, but this association did not reach statistical significance, which may be due to the small sample size and case number. Similar association was observed for everyday hair products in Black women. As such, large-scale prospective epidemiological studies with diverse populations are needed to determine the impact of PCP use on racial/ethnic disparities in asthma risk.

We used self-reported asthma status in this study, which was previously found to be valid in North America and Europe when compared to medical records ^32–36^. In particular, a study conducted in Canada found good agreement between administrative health data and self-reported asthma status, with proportions ranging from 88 to 91%, and Kappas ranging from 0.57 (95% CI: 0.52–0.63) to 0.67 (95% CI: 0.62–0.72) ^36^. Furthermore, a previous study conducted among European populations found good agreement between self-reported and registered age at adult-onset asthma among patients ^37^.

Our finding suggested that EDCs potentially have a greater impact on the respiratory health of postmenopausal women. In support of this finding, it has been reported that phthalate metabolites are associated with differences in the concentrations of sex hormones that regulate many autoimmune and inflammatory diseases, including asthma, only in postmenopausal women ^38^. Additionally, the duration of exposure may be important because older women may have had greater cumulative exposure to EDCs over their lifetime. However, we cannot discount the possibility that the non-significant association found among younger premenopausal women was due to limited sample size, as the average age at enrollment was age 55 years.

Understanding the biological pathways through which EDCs may increase the risk of asthma is important for understanding disease pathogenesis. Sex hormones play a key role in asthma pathogenesis ^39^, and the sex hormone-disrupting effects of EDCs may affect the risk of adult-onset asthma ^40^. Various phenotypes of adult onset asthma have been described in adulthood; one for example in older women is associated with obesity, more symptoms, lack of T helper 2 cell (Th2) markers, and less response to steroids ^41^. Endocrine influences may have particularly contributed to this phenotype ^42^. A direct effect of EDC exposure is suppression of inflammatory processes, which may lead to insufficient immune responses against bacteria, fungi, and viruses. Previous studies have shown that a skewed balance in Th1 / Th2 can predispose children to the development of asthma ^43^. Animal studies also showed that allergic asthmatic mice exposed to EDC had a significant increase in inflammatory cell infiltration. Triclosan has been shown to stimulate non-type 2 pro-inflammatory cytokines from immune cells, and these may be potential mediators of the late-onset, non-allergic asthma phenotype ^44^. Exposure to EDCs may also worsen asthma by elevating oxidative stress and triggering the production of calcitonin gene-related peptide (CGRP) in lung tissue ^45^. In addition, EDCs may increase the risk of asthma through epigenetic changes, including alterations of DNA methylation ^46,47^. Phthalate exposures were negatively associated with TNFα methylation, and lower methylated *TNF*α 5′CGI was associated with increased risk of asthma ^47^. Further research is needed to determine whether these mechanisms are similar in adult-onset asthma.

Our study had numerous strengths. First, our study had a large sample size and the prospective study design minimizes recall bias and reverse causation. Second, we leveraged comprehensive questionnaire data on exposure to various PCPs relevant to female U.S. populations. Here, we investigated the impact of complex mixtures of PCPs, which better reflects exposure profiles in real-world settings. This is important considering that exposure to multiple EDCs can result in synergistic, antagonistic, or cumulative effects (for compounds acting via similar pathways). Further, there is substantial toxicological evidence that mixtures are more toxic than the individual compounds ^48^.

Our investigation had some limitations. First, the impact of specific EDCs could not be investigated due to lack of information on product ingredients or brands. However, self-reported product use may reflect exposures to complex mixtures of chemicals, which may be contaminants or derivatives produced during manufacturing or storage, or may not be stated on the label by manufacturers.^9^ Second, we did not have information on duration of PCP use. Since women tend to use PCPs long-term as part of their daily or monthly routine, our findings likely reflect the effects of chronic exposure to PCPs. Further epidemiological data are needed to address changes in product use patterns over time, as well as information on exposure duration. Third, although good agreements have been reported between administrative health data and self-reported asthma status, the use of self-reported asthma status does not reflect the heterogeneity of asthma. Fourth, limitations of the latent class approach include that it is agnostic to the outcome of interest, which may obscure the associations of specific products with risk of asthma. Thus, our single-product analyses provide significant additions and identified specific products that may influence asthma risk. Fifth, the results of this study are limited to women. Gender differences have been reported in the outcomes of xenoestrogen-induced asthma ^49,50^. Given the greater and more varied use of PCP in women and the importance of estrogen in regulating biological processes in women, this population may be at greater risk of adverse health outcomes related to EDC exposure. Lastly, the analyses for Black and other minority women were underpowered; therefore, further replication in diverse populations is warranted.

The current regulations for personal care products have several limitations. The U.S, Food and Drug Administration (FDA) requires that cosmetics and personal care products be safe for consumers when used as intended, and do not contain prohibited ingredients. However, except for color additives, the FDA does not require premarket approval for cosmetics, and cosmetic companies are responsible for substantiating the safety of their products and ingredients prior to marketing. In addition, manufacturers are not required by FDA to register, file ingredient information, or report adverse reactions.^51^ The Modernization of Cosmetics Regulation Act of 2022 is the most significant expansion of the FDA’s cosmetics regulatory jurisdiction since the Federal Food, Drug, and Cosmetic Act of 1938. PCPs that are not considered cosmetics are regulated variably depending on their intended use. Our findings highlight the need for regulation of PCPs and their components and highlight the importance of providing consumers with information about their potential health effects.

Our findings provide novel insights into the relationship between commonly used PCPs, which are modifiable lifestyle factors for EDC exposure, and the risk of adult-onset asthma. If our findings are confirmed in other large-scale multi-ethnic prospective studies, they support PCPs as a potentially targetable lifestyle factor to reduce the burden of adult-onset asthma among women. Our findings also reinforce the need for regulation of PCPs and their components to reduce potential health risks, particularly adult-onset asthma.

## Data Availability

All data necessary to reproduce the current analysis are publicly available upon request as described on the Sister Study website (https://sisterstudy.niehs.nih.gov/English/data-requests.htm).

## Acknowledgments

This work was supported by intramural funding from the National Heart and Lung and Blood Institute, and Intramural Research Program of the National Institute of Health, National Institute of Environmental Health Sciences (grant number Z01ES044005). All data necessary to reproduce the current analysis are publicly available upon request as described on the Sister Study website (https://sisterstudy.niehs.nih.gov/English/data-requests.htm).

