## Supplementary tables for "Personal care product use and risk of adult-onset asthma: findings from the Sister Study"

Table S1. Latent class descriptions by product category^a^

| **Product category/class** | **Class description** |
| --- | --- |
| Beauty products |  |
| A. Infrequent users | Likely to have infrequent use of blush, eyeliner, eyeshadow, foundation, lipstick, mascara, perfume, makeup remover, artificial nails, cuticle cream, nail polish, and nail polish remover |
| B. Moderate users | Likely to have moderate use of blush, eyeliner, eyeshadow, foundation, lipstick, mascara, perfume, makeup remover, artificial nails, cuticle cream, nail polish, and nail polish remover |
| C. Frequent users | Likely to have frequent use of blush, eyeliner, eyeshadow, foundation, lipstick, mascara, perfume, makeup remover, artificial nails, cuticle cream, nail polish, and nail polish remover |
| Everyday hair products |  |
| A. Infrequent users | Likely to have infrequent use of hair conditioner, hair food, hair spray, hair gel/mousse, minoxidil, pomade, and shampoo |
| B. Moderate users | Likely to have infrequent use of hair food, minoxidil, pomade; moderate use of hair conditioner, hair spray, hair gel/mousse, and shampoo |
| C. Frequent users | Likely to have frequent use of hair conditioner, hair food, hair spray, hair gel/mousse, minoxidil, pomade, and shampoo |
| Hygiene products |  |
| A. Infrequent users | Likely to have infrequent use of shower gel, deodorant, douche, mouthwash, shaving cream, and talcum powder^b^ |
| B. Moderate users | Likely to have moderate use of shower gel, deodorant, douche, mouthwash, shaving cream, and talcum powder^b^ |
| C. Frequent users (talcum powder users) | Likely to have frequent use of shower gel, douche, shaving cream; most frequent use of deodorant, mouthwash, and talcum powder^b^ |
| Skincare products |  |
| A. Infrequent users | Likely to have infrequent use of anti-aging/wrinkle product, age spot lightener, body oil, blemish/acne product, body lotion, cleansing cream, face cream, facial mask, foot cream, hand lotion, lip moisturizer, petroleum jelly, skin lightener, and self-tanning product |
| B. Moderate users | Likely to have moderate use of anti-aging/wrinkle product, age spot lightener, body oil, blemish/acne product, body lotion, cleansing cream, face cream, facial mask, foot cream, hand lotion, lip moisturizer, petroleum jelly, skin lightener, and self-tanning product |
| C. Frequent users | Likely to have frequent use of anti-aging/wrinkle product, age spot lightener, body oil, blemish/acne product, body lotion, cleansing cream, face cream, facial mask, foot cream, hand lotion, lip moisturizer, petroleum jelly, skin lightener, and self-tanning product |

^a^Participants missing all items in each product category were excluded. Class labels and descriptions are based on likely item response probabilities for each product, but all responses for individual women may not fit these parameters; each class is described relative to the other classes in each product category (Taylor etal.2017).

^b^Refers to three different uses of talcum powder: talcum powder applied under arms, talcum powder applied vaginal area, and talcum powder applied elsewhere.

Table S2. Hazard ratios (HR) and 95% confidence intervals (CI) for the association between personal care product latent classes and adult-onset asthma risk after excluding people with asthma symptoms at baseline

|  | Infrequent users | Moderate users | Frequent users | P-trend |
| --- | --- | --- | --- | --- |
| Beauty products |  |  |  |  |
| Cases/n^a^ | 451/11,194 | 598/12,740 | 699/15,340 |  |
| HR (95%CI)^b,c^ | 1 | 1.21 (1.07, 1.37) | 1.22 (1.08, 1.38) | 0.003 |
| Everyday hair products |  |  |  |  |
| Cases/n^a^ | 137/3,648 | 965/20,996 | 644/14,604 |  |
| HR (95%CI)^b,c^ | 1 | 1.08 (0.86, 1.37) | 0.98 (0.76, 1.25) | 0.141 |
| Hygiene products |  |  |  |  |
| Cases/n^a^ | 545/14,686 | 702/14,808 | 500/9,763 |  |
| HR (95%CI)^b,c^ | 1 | 1.13 (1.004, 1.28) | 1.19 (1.05, 1.35) | 0.007 |
| Skincare products |  |  |  |  |
| Cases/n^a^ | 463/11,098 | 492/10,034 | 793/18,141 |  |
| HR (95%CI)^b,c^ | 1 | 1.18 (1.04, 1.35) | 1.16 (1.03, 1.31) | 0.024 |

^a^ Numbers of total subjects and asthma events are for women with complete data for each product class and age at date of asthma diagnosis or censoring only

^b^ Models accounted for age as the timescale.

^c^ In addition to the adjustments described in ^1^, models were adjusted for race/ethnicity (African American/Black, Hispanic/Latina non-Black, non-Hispanic White, other, or missing), education level (high school or less, some college, college and above, or missing), annual household income (<$50,000, $50,000-<$100,000, ≥$100,000), menopausal status at enrollment (premenopausal, postmenopausal, or missing), smoking status (never, past, current, or missing), secondhand smoke exposure (continuous, y), alcohol consumption (never or past, current <1 drink, current ≥1 drinks, or missing), and body mass index (continuous, kg/m^2^)

Table S3. Association between frequency of individual beauty product use in the past 12 months and adult-onset asthma risk^b^

|  | Did not use | Less than once a month | 1-3 times per month | 1-5 times per week | More than 5 times per week | P-trend |
| --- | --- | --- | --- | --- | --- | --- |
|  | HR (95%CI) | HR (95%CI) | HR (95%CI) | HR (95%CI) | HR (95%CI) |  |
| Blush or rouge | 1.00 (ref.) | 0.91 (0.76, 1.09) | 0.98 (0.81, 1.19) | 1.25 (1.09, 1.44)^a^ | 1.18 (1.03, 1.35)^a^ | <0.01 |
| Eye liner | 1.00 (ref.) | 1.13 (0.96, 1.32) | 1.08 (0.90, 1.31) | 1.17 (1.02, 1.34)^a^ | 1.06 (0.93, 1.20) | 0.18 |
| Eye shadow | 1.00 (ref.) | 1.05 (0.91, 1.21) | 1.20 (1.02, 1.41)^a^ | 1.08 (0.94 , 1.24) | 1.11 (0.97, 1.26) | 0.11 |
| Foundation makeup | 1.00 (ref.) | 1.08 (1.91, 1.29)^a^ | 1.23 (1.03, 1.48)^a^ | 1.18 (1.03, 1.35)^a^ | 1.11 (0.97, 1.25) | 0.07 |
| Lipstick | 1.00 (ref.) | 1.01 (0.83, 1.23) | 1.07 (0.88, 1.30) | 1.07 (0.91, 1.26) | 1.18 (1.01, 1.37)^a^ | 0.01 |
| Eye mascara | 1.00 (ref.) | 0.96 (0.81, 1.14) | 1.13 (0.94, 1.37) | 1.08 (0.93, 1.26) | 1.13 (0.99, 1.29) | 0.03 |
| Perfume or cologne | 1.00 (ref.) | 0.86 (0.74, 1.00) | 0.81 (0.69, 0.96) | 0.88 (0.77, 1.02) | 0.93 (0.80, 1.07) | 0.52 |
| Makeup remover | 1.00 (ref.) | 0.93 (0.79, 1.09) | 1.09 (0.90, 1.32) | 1.08 (0.92, 1.26) | 1.06 (0.91, 1.22) | 0.26 |
| Artificial nails/fill-ins someone else | 1.00 (ref.) | 1.27 (0.79, 2.06) | 0.45 (0.15, 1.40) | - | - | 0.38 |
| Artificial nails or fill-ins | 1.00 (ref.) | 1.32 (1.09, 1.59)^a^ | 1.21 (1.04, 1.42)^a^ | 3.18 (1.58, 6.38)^a^ | 2.00 (0.64, 6.23) | <0.01 |
| Cuticle cream | 1.00 (ref.) | 1.04 (0.94, 1.16) | 1.19 (1.04, 1.37)^a^ | 1.26 (0.96, 1.66) | 1.13 (0.58, 2.18) | <0.01 |
| Nail polish | 1.00 (ref.) | 1.04 (0.91, 1.20) | 1.09 (0.94, 1.26) | 1.03 (0.83, 1.28) | 1.20 (0.78, 1.86) | 0.30 |
| Nail polish remover | 1.00 (ref.) | 1.07 (0.93, 1.22) | 1.08 (0.93, 1.24) | 1.20 (0.96, 1.49) | 1.01 (0.33, 3.17) | 0.17 |

^a^ P values are less than 0.05.

^b^ Models accounted for age as the timescale. In addition, models were adjusted for race/ethnicity (African American/Black, Hispanic/Latina non-Black, non-Hispanic White, other, or missing), education level (high school or less, some college, college and above, or missing), annual household income (<$50,000, $50,000-<$100,000, ≥$100,000), menopausal status at enrollment (premenopausal, postmenopausal, or missing), smoking status (never, past, current, or missing), secondhand smoke exposure (continuous, y), alcohol consumption (never or past, current <1 drink, current ≥1 drinks, or missing), and body mass index (continuous, kg/m^2^)

Table S4. Association between frequency of individual everyday hair product use in the past 12 months and adult-onset asthma risk^b^

|  | Did not use | Less than once a month | 1-3 times per month | 1-5 times per week | More than 5 times per week | P-trend |
| --- | --- | --- | --- | --- | --- | --- |
|  | HR (95%CI) | HR (95%CI) | HR (95%CI) | HR (95%CI) | HR (95%CI) |  |
| Hair conditioner/rinse | 1.00 (ref.) | 0.82 (0.64, 1.04) | 1.05 (0.86, 1.28) | 1.10 (0.94, 1.29) | 1.01 (0.85, 1.20) | 0.23 |
| Hair food | 1.00 (ref.) | 1.02 (0.75, 1.38) | 1.25 (0.87, 1.78) | 1.57 (0.93, 2.64) | 0.82 (0.27, 2.56) | 0.15 |
| Hair spray | 1.00 (ref.) | 1.19 (1.03, 1.38)^a^ | 1.16 (0.99, 1.37) | 1.17 (1.02, 1.34)^a^ | 1.02 (0.89, 1.17) | 0.66 |
| Hair styling gel/mousse | 1.00 (ref.) | 1.13 (0.97, 1.31) | 1.06 (0.90, 1.25) | 1.17 (1.03, 1.34)^a^ | 1.02 (0.87, 1.19) | 0.30 |
| Minoxidil or Rogaine | 1.00 (ref.) | 0.37 (0.09, 1.50) | 0.61 (0.20, 1.91) | 0.87 (0.36, 2.09) | 0.74 (0.39, 1.43) | 0.19 |
| Pomade or hair grease | 1.00 (ref.) | 0.95 (0.73, 1.22) | 1.43 (1.13, 1.80)^a^ | 1.28 (1.01, 1.61)^a^ | 1.28 (0.87, 1.87) | <0.01 |
| Shampoo | 1.00 (ref.) | 1.08 (0.24, 4.97) | 0.89 (0.22, 3.62) | 0.91 (0.23, 3.64) | 0.85 (0.21, 3.40) | 0.21 |

^a^ P values are less than 0.05.

^b^ Models accounted for age as the timescale. In addition, models were adjusted for race/ethnicity (African American/Black, Hispanic/Latina non-Black, non-Hispanic White, other, or missing), education level (high school or less, some college, college and above, or missing), annual household income (<$50,000, $50,000-<$100,000, ≥$100,000), menopausal status at enrollment (premenopausal, postmenopausal, or missing), smoking status (never, past, current, or missing), secondhand smoke exposure (continuous, y), alcohol consumption (never or past, current <1 drink, current ≥1 drinks, or missing), and body mass index (continuous, kg/m^2^)

Table S5. Association between frequency of individual hygiene product use in the past 12 months and adult-onset asthma risk^b^

|  | Did not use | Less than once a month | 1-3 times per month | 1-5 times per week | More than 5 times per week | P-trend |
| --- | --- | --- | --- | --- | --- | --- |
|  | HR (95%CI) | HR (95%CI) | HR (95%CI) | HR (95%CI) | HR (95%CI) |  |
| Bath or shower gel | 1.00 (ref.) | 1.00 (0.86, 1.15) | 0.86 (0.73, 1.01) | 1.05 (0.91, 1.20) | 0.99 (0.86, 1.13) | 0.95 |
| Deodorant and/or antiperspirant | 1.00 (ref.) | 0.86 (0.52, 1.44) | 1.15 (0.77, 1.74) | 1.13 (0.83, 1.53) | 1.18 (0.90, 1.55) | 0.11 |
| Douche | 1.00 (ref.) | 0.99 (0.85, 1.16) | 1.00 (0.73, 1.35) | 1.10 (0.49, 2.45) | 0.68 (0.10, 4.83) | 0.93 |
| Mouthwash/rinse | 1.00 (ref.) | 1.10 (0.96, 1.27) | 1.14 (0.99, 1.33) | 1.12 (0.97, 1.30) | 1.13 (0.98, 1.31) | 0.09 |
| Shaving creams or gels | 1.00 (ref.) | 1.02 (0.89, 1.18) | 0.96 (0.84, 1.10) | 0.99 (0.86, 1.14) | 1.08 (0.79, 1.48) | 0.89 |
| Talcum powder under arms | 1.00 (ref.) | 1.13 (0.96, 1.34) | 0.96 (0.73, 1.26) | 0.69 (0.49, 0.96)^a^ | 1.12 (0.85, 1.48) | 0.68 |
| Talcum powder on vaginal area | 1.00 (ref.) | 0.97 (0.80, 1.18) | 1.11 (0.87, 1.42) | 1.31 (1.02, 1.68)^a^ | 1.26 (0.96, 1.66) | 0.02 |
| Talcum powder to other areas | 1.00 (ref.) | 0.98 (0.86, 1.11) | 0.99 (0.84, 1.18) | 1.13 (0.94, 1.35) | 1.09 (0.89, 1.33) | 0.27 |

^a^ P values are less than 0.05.

^b^ Models accounted for age as the timescale. In addition, models were adjusted for race/ethnicity (African American/Black, Hispanic/Latina non-Black, non-Hispanic White, other, or missing), education level (high school or less, some college, college and above, or missing), annual household income (<$50,000, $50,000-<$100,000, ≥$100,000), menopausal status at enrollment (premenopausal, postmenopausal, or missing), smoking status (never, past, current, or missing), secondhand smoke exposure (continuous, y), alcohol consumption (never or past, current <1 drink, current ≥1 drinks, or missing), and body mass index (continuous, kg/m^2^)

Table S6. Association between frequency of individual skincare product use in the past 12 months and adult-onset asthma risk^b^

|  | Did not use | Less than once a month | 1-3 times per month | 1-5 times per week | More than 5 times per week | P-trend |
| --- | --- | --- | --- | --- | --- | --- |
|  | HR (95%CI) | HR (95%CI) | HR (95%CI) | HR (95%CI) | HR (95%CI) |  |
| Anti-aging/wrinkle products | 1.00 (ref.) | 1.20 (1.03-1.41)^a^ | 1.23 (1.04-1.46)^a^ | 1.17 (1.01-1.34)^a^ | 1.13 (0.99-1.28) | 0.03 |
| Age spot lightener | 1.00 (ref.) | 1.18 (0.96-1.45) | 1.26 (0.98-1.63) | 1.06 (0.80-1.39) | 1.42 (1.08-1.87)^a^ | <0.01 |
| Baby oil/mineral-based oils | 1.00 (ref.) | 1.00 (0.86-1.15) | 0.93 (0.76-1.14) | 0.88 (0.69-1.13) | 0.86 (0.65-1.14) | 0.15 |
| Blemish/acne products | 1.00 (ref.) | 0.85 (0.72-1.00) | 1.11 (0.91-1.35) | 1.34 (1.08-1.67)^a^ | 1.09 (0.85-1.39) | 0.07 |
| Body lotions or creams | 1.00 (ref.) | 1.02 (0.76-1.36) | 1.14 (0.89-1.46) | 1.09 (0.87-1.37) | 1.16 (0.92-1.45) | 0.13 |
| Cleansing cream | 1.00 (ref.) | 0.93 (0.78-1.10) | 1.18 (1.00-1.40)^a^ | 1.07 (0.92-1.25) | 1.02(0.90-1.15) | 0.40 |
| Face creams/moisturizers | 1.00 (ref.) | 0.99 (0.76-1.30) | 1.13 (0.90-1.43) | 1.04 (0.86-1.26) | 1.02 (0.87-1.21) | 0.95 |
| Facial masks | 1.00 (ref.) | 1.09 (0.98-1.21) | 1.19 (1.00-1.41)^a^ | 0.87 (0.55-1.37) | 0.57 (0.14-2.29) | 0.13 |
| Foot creams or moisturizers | 1.00 (ref.) | 1.11 (0.97-1.28) | 1.10 (0.96-1.26) | 1.13 (0.98-1.30) | 1.05 (0.89-1.23) | 0.31 |
| Hand lotions or creams | 1.00 (ref.) | 0.66 (0.45-0.98)^a^ | 0.74 (0.53-1.05) | 0.80 (0.58-1.10) | 0.81 (0.59-1.11) | 0.44 |
| Lip moisturizers | 1.00 (ref.) | 0.88 (0.74-1.05) | 0.91 (0.77-1.08) | 0.91 (0.78-1.07) | 0.90 (0.77-1.05) | 0.38 |
| Petroleum jelly | 1.00 (ref.) | 1.01 (0.89 -1.16) | 1.12 (0.93 -1.35) | 0.93 (0.73-1.17) | 1.10 (0.88-1.37) | 0.52 |
| Skin lighteners | 1.00 (ref.) | 1.16 (0.83-1.63) | 0.90 (0.56-1.43) | 1.08 (0.71 -1.64) | 1.28 (0.83-1.97) | 0.34 |
| Self-tanning products | 1.00 (ref.) | 1.17 (1.03-1.34)^a^ | 1.07 (0.89-1.28) | 0.90 (0.68-1.18) | 1.45 (0.84-2.50) | 0.30 |

^a^ P values are less than 0.05.

^b^ Models accounted for age as the timescale. In addition, models were adjusted for race/ethnicity (African American/Black, Hispanic/Latina non-Black, non-Hispanic White, other, or missing), education level (high school or less, some college, college and above, or missing), annual household income (<$50,000, $50,000-<$100,000, ≥$100,000), menopausal status at enrollment (premenopausal, postmenopausal, or missing), smoking status (never, past, current, or missing), secondhand smoke exposure (continuous, y), alcohol consumption (never or past, current <1 drink, current ≥1 drinks, or missing), and body mass index (continuous, kg/m^2^)
